# Population-level hypertension control rate in India: A systematic review and meta-analysis of community based non-interventional studies, 2001-2020

**DOI:** 10.1101/2022.04.09.22273638

**Authors:** Shaffi Fazaludeen Koya, Zarin Pilakkadavath, Tom Wilson, Praseeda Chandran, Serin Kuriakose, Suni K Akbar, Althaf Ali

## Abstract

**Background:** Hypertension is a significant contributor to mortality in India. Adequate control of hypertension is important to prevent cardiovascular morbidity and mortality.

**Methods:** We conducted a systematic review and meta-analysis of community-based, non-interventional studies published between 2001 and 2020. We screened records from PubMed, Embase, and Web of Science databases, extracted data, and assessed risk of bias. We conducted random-effects meta-analysis to provide overall summary estimates and subgroup estimates, and mixed-effects meta-regression with sex, region, and study period as covariates. The risk of bias was assessed using modified New Castle-Ottawa scales. This study is registered with PROSPERO, CRD42021267973.

**Results:** The systematic review included 37 studies (n=170,631 hypertensive patients). Twelve studies (32%) reported poorer control rates among males than females, four studies (11%) reported poorer control rates among rural patients, while very few studies reported differences across socioeconomic variables. The overall control rate was 33.2% (n=84,485, 95% CI=27.9,38.6) with substantial heterogeneity (I2=99.1%, \chi^2= 3003.91, 95% CI=98.9,99.2; p <0.001). Unadjusted sub-group analysis showed significantly different hypertension control rates across regions (n=12,938, p=0.003) but not across study periods (n= 84,485, p=0.22), or sex (n= 81,197, p=0.22). Meta-regression showed that control rates increased by 14.7% during 2011-2020 compared to 2001-2010 (95%CI=5.8, 23.5, p=0.0021), and was 26.3% higher in the south (95%CI=12.6, 39.9, p=0.0005) and 15.9% higher in the west (95%CI=3.4, 31.4, p=0.0456) compared to the east. The control rates did not differ by sex.

**Conclusion:** Hypertension is adequately controlled only among one-third of patients in India. The control rate has improved during 2011-2020 compared to 2001-2010, but substantial differences exist across regions. Very few studies examined relevant socioeconomic factors relevant to hypertension control. India needs more studies at the community level to understand the health system and socioeconomic factors that determine uncontrolled hypertension in India.

## Introduction

Hypertension is a major modifiable risk factor for cardiovascular diseases (CVD), making it one of the major contributors of premature death and morbidity.^1,2^ The overall age adjusted prevalence of hypertension has plateaued, but the absolute number has doubled due to an increasing trend in low-middle income countries (LMICs).^3^ Globally, only 21% known hypertensive patients had their blood pressure under control.^1^ Hypertension is the most important risk factor of death and disability in India.^4,5^ The recently concluded National Family Health Survey (NFHS-5, 2019-20) reported hypertension prevalence to be 23.9% and 21.3% among men and women up from 19% and 17% respectively from the previous round.^6^

Pharmacological intervention remains the mainstay of hypertension management, and medication adherence is a cost effective way to reduce mortality and complications.^7^ Close to 80% of NCD patients in India seek medical care from the private sector, where there are no mechanisms to actively monitor drug adherence.^8^ Besides, the high out-of-pocket expenditure and lack of insurance coverage for out-patient services and drugs reduces access to anti-hypertensive medication, increasing the risk of uncontrolled hypertension.^9^

There have been no published systematic reviews or meta-analysis in the recent period, and the previous review did not explore the changes in control rates over years. This review tries to answer the following questions:

1. What is the overall hypertension control rate in India?
2. What are the sex- and region-specific estimates of control rates?
3. Whether control rate in India has improved after the launch of India’s NCD control program in 2010?

## Methods

This systematic review was performed to according to the Preferred Reporting Items for Systematic Reviews and Meta-Analyses (PRISMA) recommendations.^10^ Institutional review board approval was not required for this study since no patient identifiers were involved. The review is registered with the PROSPERO database (CRD42021267973).

### Search strategy

We searched PubMed, Web of Science, and Embase and are reported as of 31 July 2021. The search strategy (see Supplement (S1) used a combination of MeSH and non-MeSH terms for ‘hypertension’ and ‘control’. We included community-based non-interventional studies published between 1 January 2001 and 31 December 2020.

### Study eligibility

We excluded studies on secondary hypertension, interventional studies, qualitative studies, hospital-based studies, commentaries, and reviews. Studies that used convenient sampling and those that did not provide the number of known cases of hypertension were excluded.

### Data extraction

After excluding duplicates, two authors (SFK and ZP) screened all the titles and abstracts using Rayyan online collaborative systematic review platform(Figure 1).^11^ Each full text article was read by at least two authors following the inclusion criteria. Thereafter, two authors reviewed independently and extracted the following relevant information from each paper: authors, published year, study/data collection year, state, geographical area covered (rural/urban), sample size (sex-disaggregated), definitions of hypertension and control, total hypertension cases and percentage (disaggregated across sex and rural/urban), control rates (number and percentage, disaggregated numbers and percentages across sex), and reported difference in control rates across rural/urban, education levels, income status (rich/poor), and antihypertension medication status. Disagreements between reviewers were sorted out through discussions and pending discrepancies were resolved by the lead reviewer.

**Figure 1:**
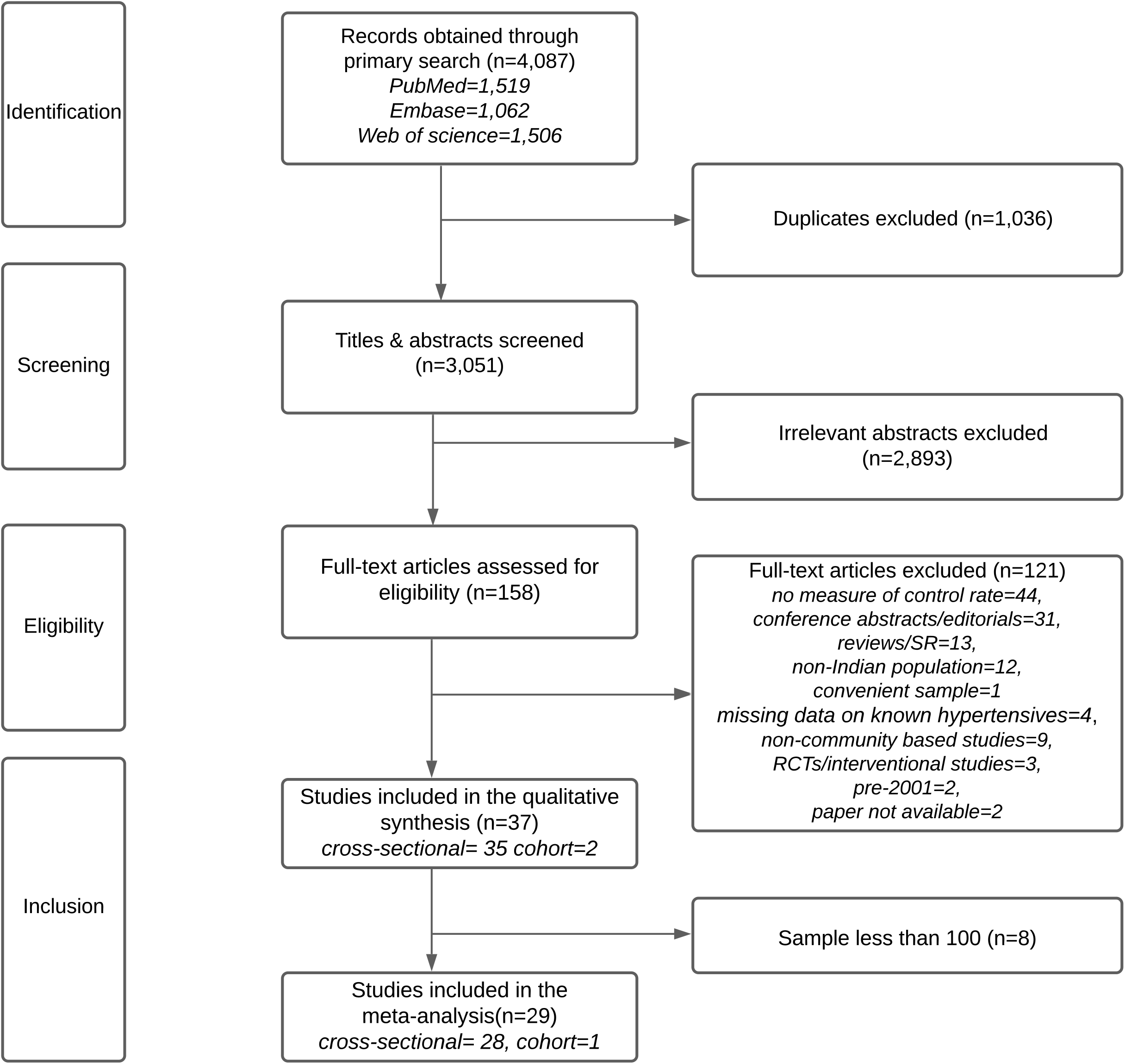
PRISMA flowchart showing study selection

We excluded 121 articles due to the following reasons: missing data, wrong article type, wrong population, wrong period, intervention studies, or full paper not available. Of the 37 articles in the review a subset of 29 articles were included in the meta-analysis (Supplement, S2) after excluding eight studies with sample size less than 100.

### Definitions used

We included known primary hypertensive adult population (18 years and above) irrespective of medication history in our denominator. We defined hypertension control as systolic blood pressure (SBP) less than 140 mmHg and diastolic blood pressure (DBP) less than 90 mmHg (JNC 7) among these known patients.^12^

### Study quality

We adapted Newcastle-Ottawa scales^13^ to assess the study quality on a scale of zero to six, across the following four criteria: selection, comparability, completeness, and statistical test. After one of the reviewers scored each paper the lead reviewer reviewed them again to decide on the score and classification. Studies that scored four or more were classified “low risk of bias” and others were classified “high risk of bias”. The median score was four; five studies got a full score of six and one study received zero. There were 19 “low risk of bias” studies and 18 “high risk of bias” studies. (Figure S3). The detailed method used for scoring and the score for each paper are shown in the supplement (S4).

### Statistical analysis

We conducted all the analysis using R software version 4.1.1 (R Core Team, 2020), and the ‘metafor’ and ‘meta’ packages were used for meta-analysis. First, we described the study characteristics using numbers and proportions and reported hypertension control rates in percentages. Second, we summarized the reported difference in control rates across sex, geography (rural/urban), education levels, income status, and by antihypertension medication status. The p-values or 95% confidence intervals were considered to decide on differences between reported rates. Finally, we did the meta-analysis and meta-regression. The summary effect size statistic for analysis- the untransformed (raw) hypertension control rates- was found to be normally distributed using Q-Q plot. Since the studies came from different regions of the country having different population characteristics, we anticipated heterogeneity and therefore decided to use random effect model *a priori*.

We used multiple methods to examine heterogeneity in our data. First, we created forest plot to visually inspect the data. Second, we looked at the total amount of systematic differences in effects across studies calculated as the between-study variance (heterogeneity, measured as τ^2^(tau-squared)) and standard deviation (τ). We used the DerSimonian-Laird estimator^14^ to calculate the heterogeneity variance (τ^2^) and Jackson method^15^ to calculate its 95% Confidence Intervals (CI) with Knapp-Hartung adjustments.^16^ Third, we estimated the I^2^ statistic (with 95% CIs)^17^ which is the ratio of observed heterogeneity (between-study variance) and the total observed variance (sum of within-study variance due to sampling error and between-study variance). Finally, we conducted a formal x2 test with a Cochran’s Q statistic, to test if all studies share the common effect size.^18^ All statistical tests were two sided and p-value was fixed at 0.05.

## Results

### Study characteristics

Table 1 shows the overall features of the studies included in the review. The systematic review includes 37 studies (35 cross-sectional and two cohort studies).^19–54^ The total sample was 870,659 (80% females) including 170,631 hypertensive patients. The mean hypertension prevalence across studies was 35.6% (SD= 14.6) which did not vary between males and females.

**Table 1:**
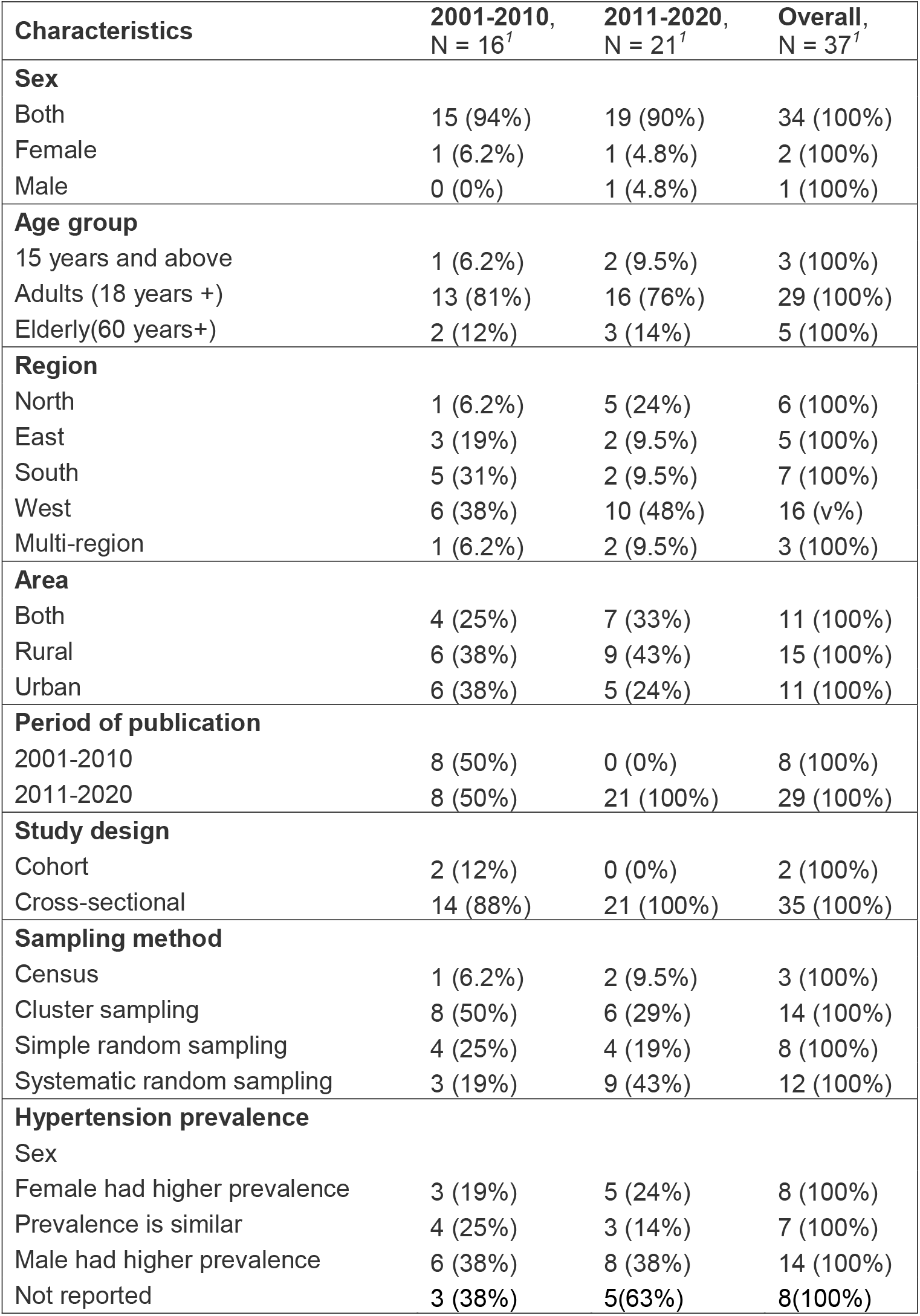
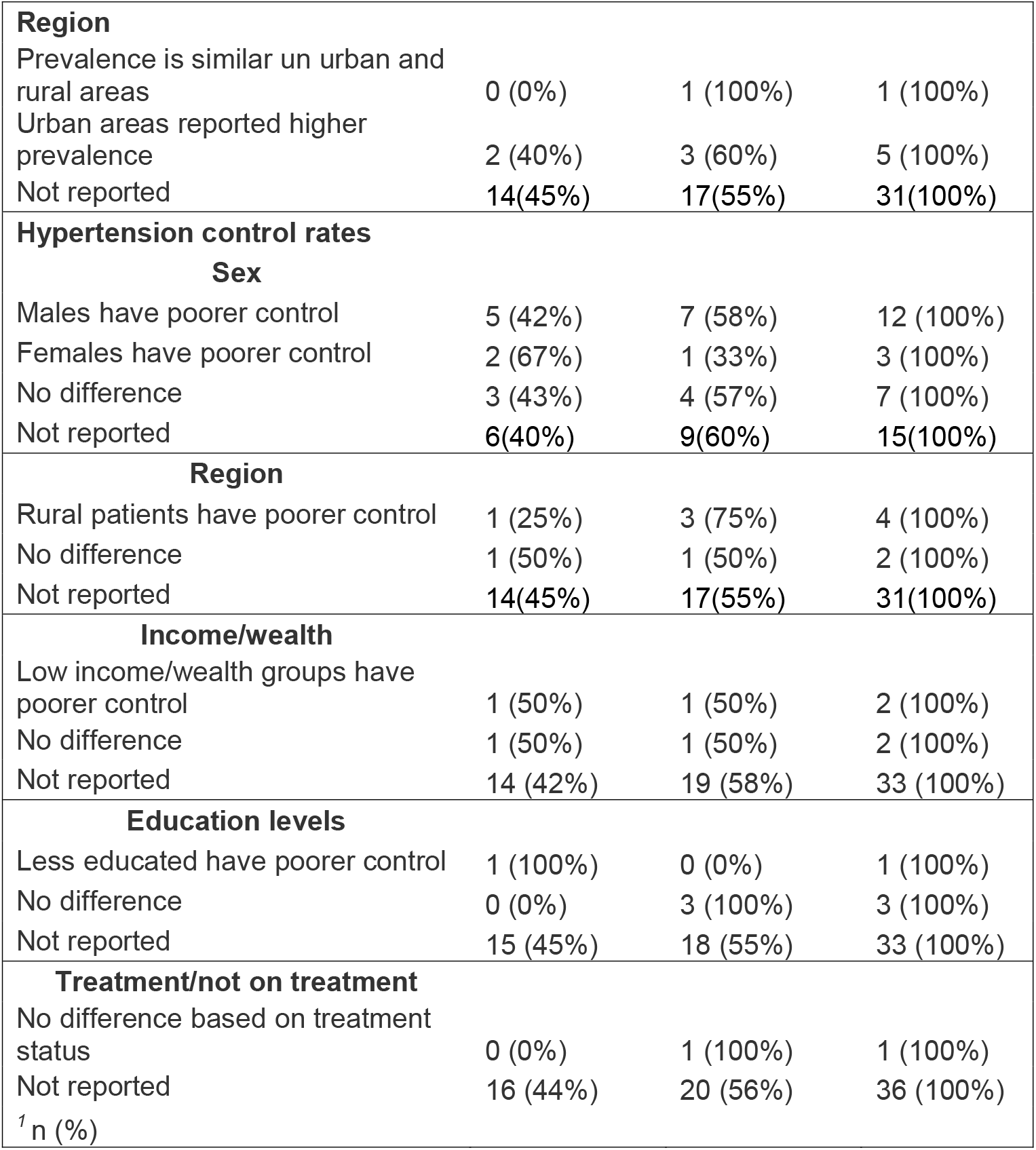
Summary characteristics of studies included in the systematic review.

| Characteristics | 2001-2010,<br>N = 16<sup>1</sup> | 2011-2020,<br>N = 21<sup>1</sup> | Overall,<br>N = 37<sup>1</sup> |
| --- | --- | --- | --- |
| **Sex** |  |  |  |
| Both | 15 (94%) | 19 (90%) | 34 (100%) |
| Female | 1 (6.2%) | 1 (4.8%) | 2 (100%) |
| Male | 0 (0%) | 1 (4.8%) | 1 (100%) |
| **Age group** |  |  |  |
| 15 years and above | 1 (6.2%) | 2 (9.5%) | 3 (100%) |
| Adults (18 years +) | 13 (81%) | 16 (76%) | 29 (100%) |
| Elderly(60 years+) | 2 (12%) | 3 (14%) | 5 (100%) |
| **Region** |  |  |  |
| North | 1 (6.2%) | 5 (24%) | 6 (100%) |
| East | 3 (19%) | 2 (9.5%) | 5 (100%) |
| South | 5 (31%) | 2 (9.5%) | 7 (100%) |
| West | 6 (38%) | 10 (48%) | 16 (v%) |
| Multi-region | 1 (6.2%) | 2 (9.5%) | 3 (100%) |
| **Area** |  |  |  |
| Both | 4 (25%) | 7 (33%) | 11 (100%) |
| Rural | 6 (38%) | 9 (43%) | 15 (100%) |
| Urban | 6 (38%) | 5 (24%) | 11 (100%) |
| **Period of publication** |  |  |  |
| 2001-2010 | 8 (50%) | 0 (0%) | 8 (100%) |
| 2011-2020 | 8 (50%) | 21 (100%) | 29 (100%) |
| **Study design** |  |  |  |
| Cohort | 2 (12%) | 0 (0%) | 2 (100%) |
| Cross-sectional | 14 (88%) | 21 (100%) | 35 (100%) |
| **Sampling method** |  |  |  |
| Census | 1 (6.2%) | 2 (9.5%) | 3 (100%) |
| Cluster sampling | 8 (50%) | 6 (29%) | 14 (100%) |
| Simple random sampling | 4 (25%) | 4 (19%) | 8 (100%) |
| Systematic random sampling | 3 (19%) | 9 (43%) | 12 (100%) |
| **Hypertension prevalence** |  |  |  |
| Sex |  |  |  |
| Female had higher prevalence | 3 (19%) | 5 (24%) | 8 (100%) |
| Prevalence is similar | 4 (25%) | 3 (14%) | 7 (100%) |
| Male had higher prevalence | 6 (38%) | 8 (38%) | 14 (100%) |
| Not reported | 3 (38%) | 5(63%) | 8(100%) |
| **Region** |  |  |  |
| Prevalence is similar un urban and rural areas | 0 (0%) | 1 (100%) | 1 (100%) |
| Urban areas reported higher prevalence | 2 (40%) | 3 (60%) | 5 (100%) |
| Not reported | 14(45%) | 17(55%) | 31(100%) |
| **Hypertension control rates** |  |  |  |
| **Sex** |  |  |  |
| Males have poorer control | 5 (42%) | 7 (58%) | 12 (100%) |
| Females have poorer control | 2 (67%) | 1 (33%) | 3 (100%) |
| No difference | 3 (43%) | 4 (57%) | 7 (100%) |
| Not reported | 6(40%) | 9(60%) | 15(100%) |
| **Region** |  |  |  |
| Rural patients have poorer control | 1 (25%) | 3 (75%) | 4 (100%) |
| No difference | 1 (50%) | 1 (50%) | 2 (100%) |
| Not reported | 14(45%) | 17(55%) | 31(100%) |
| **Income/wealth** |  |  |  |
| Low income/wealth groups have poorer control | 1 (50%) | 1 (50%) | 2 (100%) |
| No difference | 1 (50%) | 1 (50%) | 2 (100%) |
| Not reported | 14 (42%) | 19 (58%) | 33 (100%) |
| **Education levels** |  |  |  |
| Less educated have poorer control | 1 (100%) | 0 (0%) | 1 (100%) |
| No difference | 0 (0%) | 3 (100%) | 3 (100%) |
| Not reported | 15 (45%) | 18 (55%) | 33 (100%) |
| **Treatment/not on treatment** |  |  |  |
| No difference based on treatment status | 0 (0%) | 1 (100%) | 1 (100%) |
| Not reported | 16 (44%) | 20 (56%) | 36 (100%) |
| <sup>1</sup> n (%) |  |  |  |

Sixteen studies (43%) reported data for the period 2001-2010 while the remaining 21 studies had data for the period 2011-2020. Forty one percent studies had data only from rural areas, and five studies (14%) reported a higher prevalence of hypertension in urban areas compared to rural areas. 16 studies (43%) were from southern states in India, most studies (n=34, 92%) had both males and females, and fourteen (38%) studies reported a higher prevalence of hypertension in males.

### Reporting of control rates

There were 12 studies (32%) that reported poorer control rates among males than females ^20,32,33,36,41,43,45,49–51,53,54^ while only three studies showed females to have a poorer control rate than males.^24,31,44^ Four studies (11%) reported poorer control rates among rural patients ^43,45,47,54^ while two studies showed there was no difference in control rate between rural and urban patients.^48,51^ Two studies showed poorer control in the low socioeconomic group.^26,54^ Only one study reported on difference in control rates based on medication status which showed no difference between the groups.^29^ One study^50^ showed poorer control in the less educated group while three studies found no difference across educational levels.^46,48,54^

### Meta-analysis

We used random effects model to calculate the summary effect size, i.e., the weighted average of the observed control rates in 29 studies. The inverse of the total variance of the study was used to weigh each study. The output revealed that τ^2^ is 0.02 (95% CI=0.01, 0.05), τ= 12.7% (95% CI= 9.9,22.2), I^2^ is 99.1% (95% CI=98.9,99.2), and the Q-statistic (df=28) is 3003.9 (p<0.0001), all of which suggested high heterogeneity in the effect sizes. To identify outliers and influential studies causing heterogeneity, we used a diagnostic Baujat plot (Supplement S5) which showed two studies with studentized residuals (z-values) greater than two.^28,45^ To further investigate, we performed a set of leave-one-out diagnostic tests (Supplement S6) to calculate the summary values of hypertension control rates by excluding one study each at a time from the analysis. However, the results and subsequent visualization or residuals (Supplement S7) did not show any significant difference in control rates with the exclusion of the two studies. So, we decided against removing any studies from the model.

### Overall hypertension control rate

The overall random effects model with Hartung-Knapp adjustment shows that the mean rate of hypertension control in India during 2001-2011 was 33.2% (95% CI= 27.9, 38.6). (Figure 2) In comparison, a post-hoc estimate of the fixed effect model shows a pooled control rate of 17.0% (95% CI=16.8%, 17.2%) lower than our estimates using random effects model. The wide difference between the models also substantiates our decision to use random-effects model. Our 95% prediction interval 6.7% - 59.8% is wide reflecting high levels of heterogeneity.

**Figure 2:**
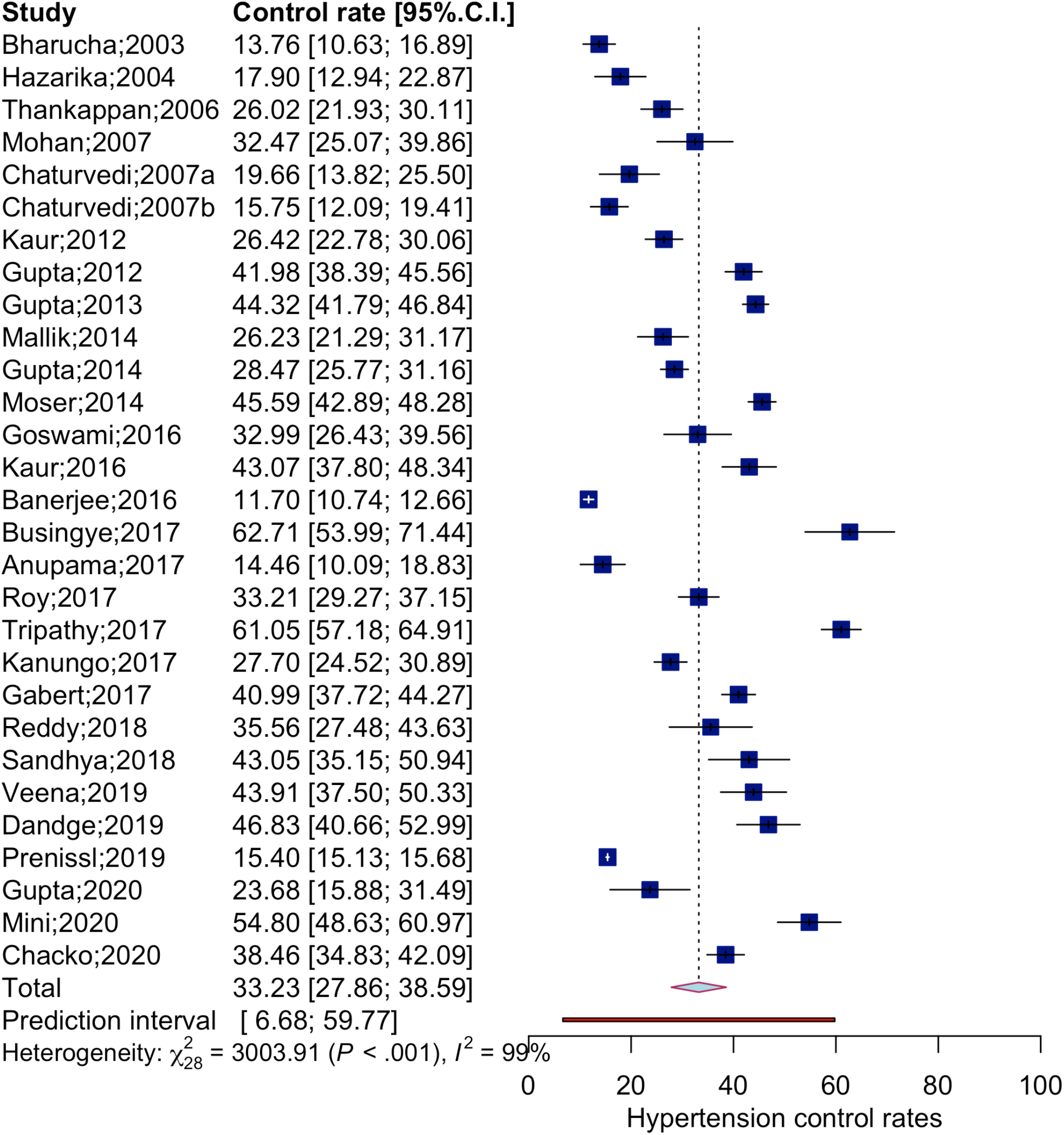
Forest plot showing the unadjusted hypertension control rates

### Subgroup analysis

We conducted subgroup analysis to understand the heterogeneity and the results are summarized as forest plot (Figure 3) and table (Supplement S8). To test if the hypertension rates have improved over time, we compared studies conducted in the first ten years (2001-2010) with studies conducted in the second ten years (2011-2020). We computed the summary effects for each subgroup under the random effects model. Since systematic reasons like differences in population across states can still produce different values of the within-group τ^2^ values, we applied separate estimates of τ^2^ for each subgroup, effectively resulting in an independent meta-analysis of the subgroups. We found that the control rates have improved over the years (35.8% in 2011-2020 versus 29.6% in 2001-2010), but the improvement was not statistically significant (p=0.22), and there was significant heterogeneity (τ^2^ = 0.01, p<0.001).

**Figure 3:**
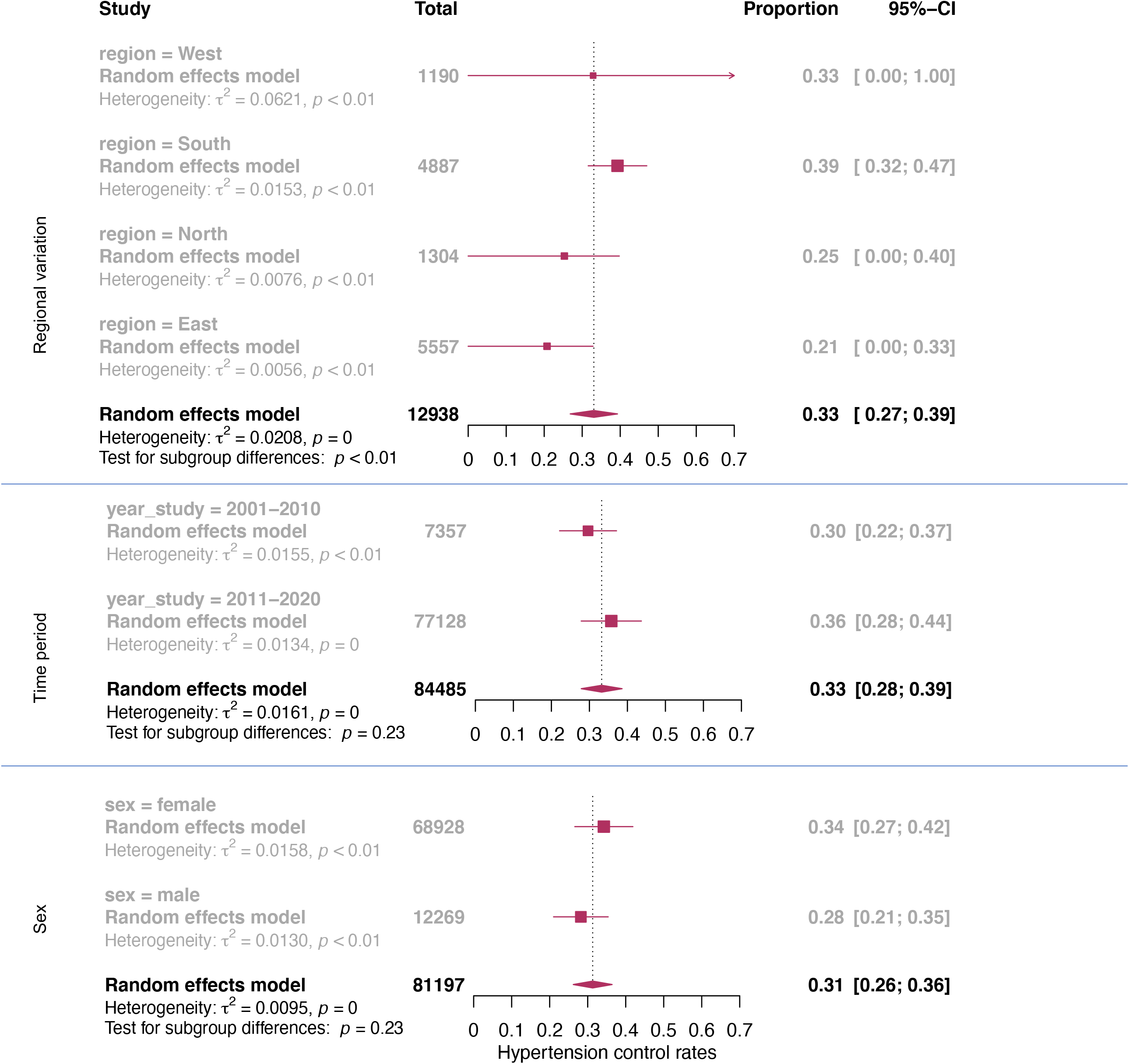
Forest plot showing sub-group analysis across region, study period and sex.

We conducted meta-analysis of 81,197 patients (68,928 females and 12,269 males) pooled from 20 studies with sex-segregated data. The results showed that females had better control rates than males, but the difference was not statistically significant (34.2% [95% CI=26.6, 41.9] for females versus 28.2% [95%CI=21.0, 35.4] for males). Substantial heterogeneity remained (τ^2^ = 0.01, p< 0.001). The control rates of females improved by 11% points between 2001-2010 and 2011-2020 whereas the control rates for males improved only by 7% points during the same period. However, these changes were not statistically significant.

In the subgroup analysis for regions, we avoided studies with data from multiple regions and analyzed 12,938 known hypertensive patients from 24 studies (13 studies from south, 4 studies each from north and east, and 3 from west). Control rates were significantly different across regions (p= 0.003). The south (39.3%) and west (32.9%) regions reported higher control rates compared to the north (25.8%) and the east (20.7%).

#### Meta-regression

To control rate for differences in region, period of study, and sex, we conducted a mixed-effects meta-regression^55,56^ using the model equation:

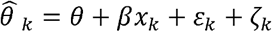

where 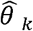 is the observed effect-size (hypertension control rate) with *k* studies, *θ* is the intercept, *β* is the regression coefficient for the variable *x, ε*_*k*_ is the sampling error through which the effect size of a study deviates from its true effect, and *ζ*_*k*_ is the error arising due to heterogeneity. We built a multiple regression model with the following variables: region(north, east, south, west), period of study (2001-’10 vs 2011-’20), and sex(male, female), after excluding studies involving multiple regions. (Table 2). The model accounted for 52% of heterogeneity and showed that control rate did not differ across sex when controlled for region and period of study. The model also showed that when adjusted for regional and sex differences, the rate of control improved by 14.7% points from 2001-2010 to 2011-2020 (p< 0.01). Southern region reported 26.3% (95% CI= 12.6, 39.9, P= 0.0005) and western region reported 15.9%(95% CI=3.4, 31.4, p= 0.0456) higher control rate compared to the eastern region.

**Table 2:** Meta-regression model: hypertension control rates in India, 2001-2020.

|  | Effect estimate | 95% CI | Standard error | t-value | p- value |
| --- | --- | --- | --- | --- | --- |
| Intercept | 9.2 | -4.3, 22.7 | 6.6 | 1.39 | 0.1743 |
| Region: North | 10.1 | -4.6, 24.7 | 7.1 | 1.41 | 0.1687 |
| Region: South | 26.3 | 12.6, 39.9*** | 6.6 | 3.96 | 0.0005 |
| Region: West | 15.9 | 3.4, 31.4* | 7.5 | 2.10 | 0.0456 |
| Period: 2011-2020 | 14.7 | 5.8, 23.5** | 4.3 | 3.43 | 0.0021 |
| Sex: Male | -4.7 | -13.5, 4.0 | 4.3 | -1.11 | 0.279 |

Significance codes: '\*\*\*' 0.001 '\*\*' 0.01 '\*' 0.05

#### Sensitivity analysis

We conducted two sensitivity analyses. The first analysis was done by avoiding four studies with only elderly population. In the second sensitivity analysis, we included only “low risk of bias” studies. The resultant models did not differ in overall control rates from the original model.

#### Publication bias

With prevalence as the outcome measure we do not expect publication bias arising from study design related significance level.^57^ The funnel plot asymmetry (Supplement, S9) and the subsequent Egger’s^58^ regression test result (t=5.6, p< 0.0001) may not be explained by publication bias but rather by the high level of heterogeneity and quality of studies themselves.

## Discussion

To our understanding this is the first meta-analysis that examined the changes in population-level hypertension control rates in India over the years. We compared the control rates in one decade preceding and one decade succeeding the launch of India’s national program for NCDs. There are four key findings from our study.

First, only one-third of known hypertensive patients in India have adequate blood pressure control despite the launch of a national program to control NCD in 2010. The only previously published meta-analysis of community-level hypertension control in India with data from 1950 to 2013 showed a control rate of 10.7% for rural India and 20.2% for urban India.^59^ Though the control rate that we report is substantially higher, the low rate of 33.2% is still a concern. This is especially true considering that only 50% of patients in 15–49-year age group in India knew their hypertension status as per the NFHS-4 data (2015-16).^54^ India’s NCD program needs serious evaluation to see how far is it meeting its public health objectives to control hypertension.^60^ Interrupted supply of medicines, inadequate health education and low health literacy can have a synergistic effect leading to incomplete treatment or non-compliance. India also started a multi-partner initiative, the India hypertension control initiative in 2017, to strengthen the public health measures to control hypertension. A recent study analyzing the initial cohort from four Indian states showed significant improvement in blood pressure control (59.8% in follow up versus 26.3% at baseline), more so in the primary care settings that shows that better blood pressure control can be achieved through scalable public health programs.^61^ Comparing with recent literature, a recent cross-sectional study of 1.1 million adults across 44 LMICs including India showed that the control rate for hypertension was only 10.3%.^62^ A systematic review and meta-analysis from Nepal showed a hypertension control rate of 38% among treated hypertensives with only marginal improvement over years.^63^ The most recent data from Pakistan shows that only half of diagnosed hypertensive patients are treated and only 12.5% are controlled.^64^

Second, significant regional differences exist in the hypertension control, even when limited by the fewer number of studies in west and north India compared to south. South India showed better control rates after adjusting for sex and study period. Kerala and Tamil Nadu reported the highest rates of control, after excluding the very high rates reported by one study each from Punjab and Andhra Pradesh. The difference in health system capacity to detect and treat hypertension varies across the country as much as the level of awareness about the disease, its prevention, and control vary. Treatment adherence and access to medicine are key determinants of adequate control. Veena et. al reported that among those with controlled hypertension, 23.7% subjects monitored blood pressure 2-4 times a year while 67.30% never monitored their blood pressure.^35^ Adherence to medications was examined in only one study^46^ in our review that showed significant association with control rate. In addition, we found only one study^29^ (conducted among elderly) that compared control status based on medication status, while no study was found to examine the access to antihypertensive medicines. A recent study had shown that low availability of generic medicines in public and private sector and high costs are major barriers to antihypertensive control including in India.^65^ Another study reported that around 70% of the estimated proportion of adults with hypertension did not receive antihypertensive drugs in 2018.^66^

Third, very few studies reported lifestyle and risk factors associated with poor control rates. Among them, Tripathy et. al^45^ reported that uncontrolled hypertension was more frequent among obese patients, patients with sedentary lifestyle, and diabetic patients. Thankappan et. al^41^ also found poor blood pressure control among diabetics and obese patients. Diet and smoking were reported as predictors in one study^46^ while greater per cent body fat was the only factor reported in another,^28^ while good family support to self-care was reported by a third study.^46^

Finally, very few studies had data on key social determinants of hypertension control like income, wealth, and caste. Data on income or wealth and education were unavailable in 89% of studies, while no studies had data on caste differences on hypertension control. A recent study (not included in our review) showed 13 percent point gap in control rate between the rich and the poor and clear disadvantage for scheduled castes, tribes and backward communities.^67^ The previous meta-analysis from India reported significant differences in rural and urban on awareness and control levels while no significant difference was noted for percentage treated.^61^ In our review we found two studies that reported no difference between urban and rural population while four studies reported rural populations to have poorer control.

### Limitations

We included only studies published until 31 December 2020, and as such we would have missed studies that have been published afterwards. Our study did not explain differences across age groups, as we were limited by the data availability in the reviewed papers.

## Conclusion

India needs far more studies at the community level to understand the epidemiology of hypertension control, especially in north and west India. Well-designed studies that ensure quality of data will help us to better understand the differences in control rates across regions. Studies should examine relevant health-system, socio-economic, and lifestyle factors that determine adequate control levels so that policies and programs can be designed to specifically address the key determinants of uncontrolled hypertension in India.

## Data Availability

All the data used in the analysis came from the articles we reviewed, and the articles are included in the reference.

## Acknowledgments

None

