## Supplement file for "Population-level hypertension control rate in India: A systematic review and meta-analysis of community based non-interventional studies, 2001-2020"

**S1: Search strategy**

**PubMed**

("Hypertension"[Mesh] OR "hypertension"[Text Word] OR "blood pressure"[Text Word] OR "raised blood pressure"[Text Word] OR "elevated blood pressure"[Text Word] OR "systolic blood pressure"[Text Word] OR "diastolic blood pressure"[Text Word] OR "SBP"[Text Word] OR "DBP"[Text Word] OR "isolated systolic blood pressure"[Text Word] OR "high BP"[Text Word] OR "BP"[Text Word] OR "raised BP"[Text Word] OR "elevated BP"[Text Word]) AND 2001/01/01:2020/12/31[Date - Publication] AND ("India"[Text Word] OR "Indian"[Text Word]) AND "control"[Text Word]

**Web of science**

(('hypertension'/exp OR hypertension OR 'blood pressure' OR 'raised blood pressure' OR 'elevated blood pressure' OR 'systolic blood pressure' OR 'diastolic blood pressure' OR 'sbp' OR 'dbp' OR 'isolated systolic blood pressure' OR 'high bp' OR 'bp' OR 'raised bp' OR 'elevated bp') AND ('indian' OR 'india' ) AND ('control')) AND [1-1-2001]/sd NOT [1-1-2021]/sd AND [2001-2020]/py

**Embase**

((('hypertension':ab,ti OR hypertension:ab,ti OR 'blood pressure':ab,ti OR 'raised blood pressure':ab,ti OR 'elevated blood pressure':ab,ti OR 'systolic blood pressure':ab,ti OR 'diastolic blood pressure':ab,ti OR 'sbp':ab,ti OR 'dbp':ab,ti OR 'isolated systolic blood pressure':ab,ti OR 'high bp':ab,ti OR 'bp':ab,ti OR 'raised bp':ab,ti OR 'elevated bp':ab,ti) AND ('indian':ab,ti OR india:ab,ti) AND 'control':ab,ti) AND (2001:py OR 2002:py OR 2003:py OR 2004:py OR 2005:py OR 2006:py OR 2007:py OR 2008:py OR 2009:py OR 2010:py OR 2011:py OR 2012:py OR 2013:py OR 2014:py OR 2015:py OR 2016:py OR 2017:py OR 2018:py OR 2019:py OR 2020:py)) AND [embase]/lim NOT ([embase]/lim AND [medline]/lim)

**S2: List of studies included in the systematic review**

| Author; year of publication | Study year | Study design | State | Region | Area | Sex | Sampling method | HTN prevalence | Known prev. | Male/female difference | HTN control rate | Male/female difference in control rate |
| --- | --- | --- | --- | --- | --- | --- | --- | --- | --- | --- | --- | --- |
| Mallik;2014 | 2011 | CS | West Bengal | E | R | both | census | 41.9 | 40.6 | u/a | 35.4 | u/a |
| Veena;2019 | 2017 | CS | Pondicherry | S | U | both | census | ## | u/a | u/a | 43.9 | u/a |
| Bharucha;2003 | 2001 | CS | Maharashtra | W | U | both | Cl | 36.4 | 52.3 | u/a | 13.9 | u/a |
| Kaur;2016 | 2009 | C | Tamil Nadu | S | R | both | SRS | 22.9 | 26.4 | u/a | 43.1 | u/a |
| Prayag;2017 | 2015 | CS | Karnataka | S | U | both | Sy.RS | 38.3 | 53.1 | f>m | 49.2 | u/a |
| Misra;2014 | 2011 | CS | Assam | E | R | both | Cl | 26 | 23.5 | f=m | 10.0 | u/a |
| Anupama;2017 | 2015 | CS | Karnataka | S | R | both | Cl | 30.4 | 27.4 | f=m | 14.4 | u/a |
| Hazarika;2004 | 2003 | CS | Assam | E | R | both | SRS | 33.3 | 21.6 | f=m | 17.9 | u/a |
| Bhardwaj;2010 | 2009 | CS | Himachal Pradesh | N | R | both | Cl | 35.9 | 22 | m>f | 20.2 | u/a |
| Yadav;2008 | 2007 | CS | Delhi | N | U | both | SRS | 39.5 | 51.7 | m>f | 28.3 | u/a |
| Dandge;2019 | 2017 | CS | Telangana | S | R | both | SRS | 23.6 | 61 | m>f | 47.0 | u/a |
| Kanungo;2017 | 2014 | CS | West Bengal | E | R&U | both | Sy.RS | 26 | 16.1 | m>f | 27.7 | u/a |
| Karmakar;2018 | 2014 | CS | West Bengal | E | R | both | Sy.RS | 26.1 | 48.2 | f>m | 18.3 | f ↓ |
| Kaur;2012 | 2007 | CS | Tamil Nadu | S | R | both | SRS | 21.5 | 25.1 | f=m | 26.4 | f ↓ |
| Mohan;2007 | 2006 | CS | Tamil Nadu | S | U | both | Sy.RS | 20 | 32.8 | m>f | 32.5 | f ↓ |
| Gupta;2014 | 2010 | CS | Multiple | M | U | both | Cl | 31.3 | 55.3 | f>m | 28.2 | m ↓ |
| Goswami;2016 | 2015 | CS | New Delhi | N | U | both | Cl | 67.2 | 41.2 | f>m | 32.9 | m ↓ |
| Moser;2014 | 2007 | CS | Kerala | S | R&U | both | Cl | 32 | 41 | f>m | 42.7 | m ↓ |
| Thankappan;2006 | 2001 | CS | Kerala | S | R | both | Cl | 36.7 | 24.4 | f>m | 26.0 | m ↓ |
| Gabert;2017 | 2013 | CS | Multiple | M | R&U | both | Sy.RS | 32.2 | 37.4 | f>m | 42.0 | m ↓ |
| Chaturvedi;2007a | 2006 | CS | Delhi | N | R&U | both | Cl | 27.5 | 53.3 | f=m | 19.7 | m ↓ |
| Kusuma;2013 | 2007 | CS | Delhi | N | U | both | census | 18.3 | 41 | m>f | 14.7 | m ↓ |
| Prenissl;2019 | 2016 | CS | All India | M | R&U | both | Cl | 17.8 | 44.7 | m>f | 17.7 | m ↓ |
| Roy;2017 | 2012 | CS | New Delhi | N | R&U | both | Cl | 35.7 | 38.7 | m>f | 33.1 | m ↓ |
| Banerjee;2016 | 2015 | CS | West Bengal | E | U | both | SRS | 42 | 54.4 | m>f | 21.3 | m ↓ |
| Mini;2020 | 2018 | CS | Kerala | S | R&U | both | Sy.RS | 14.6 | 62.2 | m>f | 54.8 | m ↓ |
| Tripathy;2017 | 2015 | CS | Punjab | W | R&U | both | Sy.RS | 40.1 | 30.1 | m>f | 61.0 | m ↓ |
| Chacko;2020 | 2019 | CS | Kerala | S | R&U | both | Cl | ## | u/a | u/a | 38.4 | nd |
| Reddy;2018 | 2017 | CS | Telangana | S | U | both | SRS | 83.5 | 80.2 | f>m | 46.2 | nd |
| Chaturvedi;2007b | 2006 | CS | Delhi | N | R&U | both | Cl | 63.8 | 54 | f=m | 15.7 | nd |
| Gupta;2020 | 2018 | CS | Haryana | W | R | both | SRS | 50.3 | 58.8 | f=m | 23.7 | nd |
| Gupta;2013 | 2010 | CS | Multiple | M | U | both | Cl | 42.8 | 56.95 | m>f | 44.3 | nd |
| Sathish;2012 | 2006 | C | Kerala | S | R | both | Sy.RS | ## | u/a | m>f | 6.7 | nd |
| Busingye;2017 | 2014 | CS | Andhra Pradesh | S | R | both | Sy.RS | ## | 42.6 | m>f | 62.7 | nd |
| Gupta;2012 | 2007 | CS | All India | M | R&U | f | Sy.RS | 39.2 | 42.8 | n/a | 42.0 | n/a |
| Begam;2016 | 2013 | CS | Kerala | S | R | m | Sy.RS | 45 | 47.9 | n/a | 50.6 | n/a |
| Sandhya;2018 | 2014 | CS | Kerala | S | R | f | Sy.RS | 32.1 | 67.7 | n/a | 43.0 | n/a |

*HTN: Hypertension, CS: cross-sectional study, C: Cohort, S: South, N: North, E: East, W: West, M- Multiple, R: rural, U: urban, m: male, f: female, Cl: Cluster sampling, SRS: Simple random sampling, Sy.RS: Systematic random sampling, u/a: data unavailable, n/a: not applicable, f* ↓: females have poorer control rate, *m* ↓: males have poorer control rate, nd: no difference

**S3: Risk of bias- summary graph**

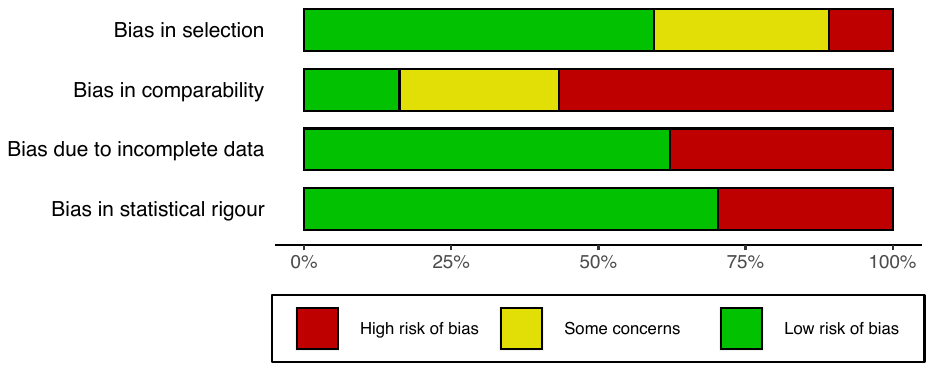

**S4: Risk of bias assessment using modified New Castle-Ottawa scales- scores of individual studies**

| Study | Selection^#^ | Comparability^$^ | Completeness^%^ | Statistics^*^ | Score [/6] | Risk of Bias |
| --- | --- | --- | --- | --- | --- | --- |
| Misra;2014 | 2 | 0 | 1 | 1 | 4 | low |
| Kusuma;2013 | 2 | 0 | 0 | 0 | 2 | high |
| Yadav;2008 | 0 | 0 | 1 | 0 | 1 | high |
| Prayag;2017 | 2 | 0 | 1 | 1 | 4 | low |
| Sathish;2012 | 1 | 1 | 1 | 1 | 4 | low |
| Karmakar;2018 | 2 | 0 | 0 | 0 | 2 | high |
| Begam;2016 | 1 | 0 | 1 | 1 | 3 | high |
| Bhardwaj;2010 | 2 | 0 | 1 | 0 | 3 | high |
| Gupta;2020 | 1 | 0 | 1 | 1 | 3 | high |
| Busingye;2017 | 1 | 1 | 0 | 1 | 3 | high |
| Reddy;2018 | 0 | 0 | 0 | 0 | 0 | high |
| Sandhya;2018 | 1 | 0 | 0 | 1 | 2 | high |
| Mohan;2007 | 2 | 0 | 1 | 1 | 4 | low |
| Chaturvedi;2007a | 1 | 0 | 0 | 0 | 1 | high |
| Goswami;2016 | 2 | 0 | 1 | 1 | 4 | low |
| Hazarika;2004 | 2 | 2 | 1 | 1 | 6 | low |
| Veena;2019 | 1 | 0 | 0 | 0 | 1 | high |
| Mini;2020 | 2 | 0 | 0 | 1 | 3 | high |
| Anupama;2017 | 2 | 1 | 1 | 1 | 5 | low |
| Dandge;2019 | 2 | 0 | 0 | 1 | 3 | high |
| Mallik;2014 | 1 | 0 | 1 | 1 | 3 | high |
| Kaur;2016 | 0 | 0 | 1 | 0 | 1 | high |
| Chaturvedi;2007b | 1 | 0 | 0 | 0 | 1 | high |
| Thankappan;2006 | 2 | 1 | 1 | 1 | 5 | low |
| Bharucha;2003 | 0 | 0 | 1 | 0 | 1 | high |
| Roy;2017 | 2 | 2 | 1 | 1 | 6 | low |
| Kaur;2012 | 2 | 1 | 1 | 1 | 5 | low |
| Tripathy;2017 | 2 | 2 | 1 | 1 | 6 | low |
| Chacko;2020 | 1 | 1 | 1 | 1 | 4 | low |
| Gupta;2012 | 1 | 1 | 0 | 1 | 3 | high |
| Kanungo;2017 | 2 | 1 | 0 | 1 | 4 | low |
| Gabert;2017 | 2 | 2 | 0 | 0 | 4 | low |
| Gupta;2015 | 2 | 2 | 1 | 1 | 6 | low |
| Moser;2014 | 2 | 1 | 1 | 1 | 5 | low |
| Gupta;2013 | 2 | 2 | 1 | 1 | 6 | low |
| Banerjee;2016 | 2 | 0 | 1 | 1 | 4 | low |
| Prenissl;2019 | 2 | 1 | 0 | 1 | 4 | low |

\# **Selection**: One point each if the study used a sample representative of the average hypertensive population in the community or if the sample size was pre-calculated or was justified and satisfactory.

$ **Comparability**: If the study accounted for at least one confounder like a demographic characteristic, one point was given, and if at least one more relevant confounder (like smoking, obesity, increased lipids, alcohol etc.) was accounted for, one more point was given.

% **Completeness**: If the response rate (cross-sectional) or follow-up (cohort) was satisfactory one point was given.

* **Statistics**: If the statistical test used to analyze the data is clearly described and appropriate and the measurement of the association is presented including confidence intervals and the probability level, then one point was given.

**S5: Diagnostic Bajaut plot**

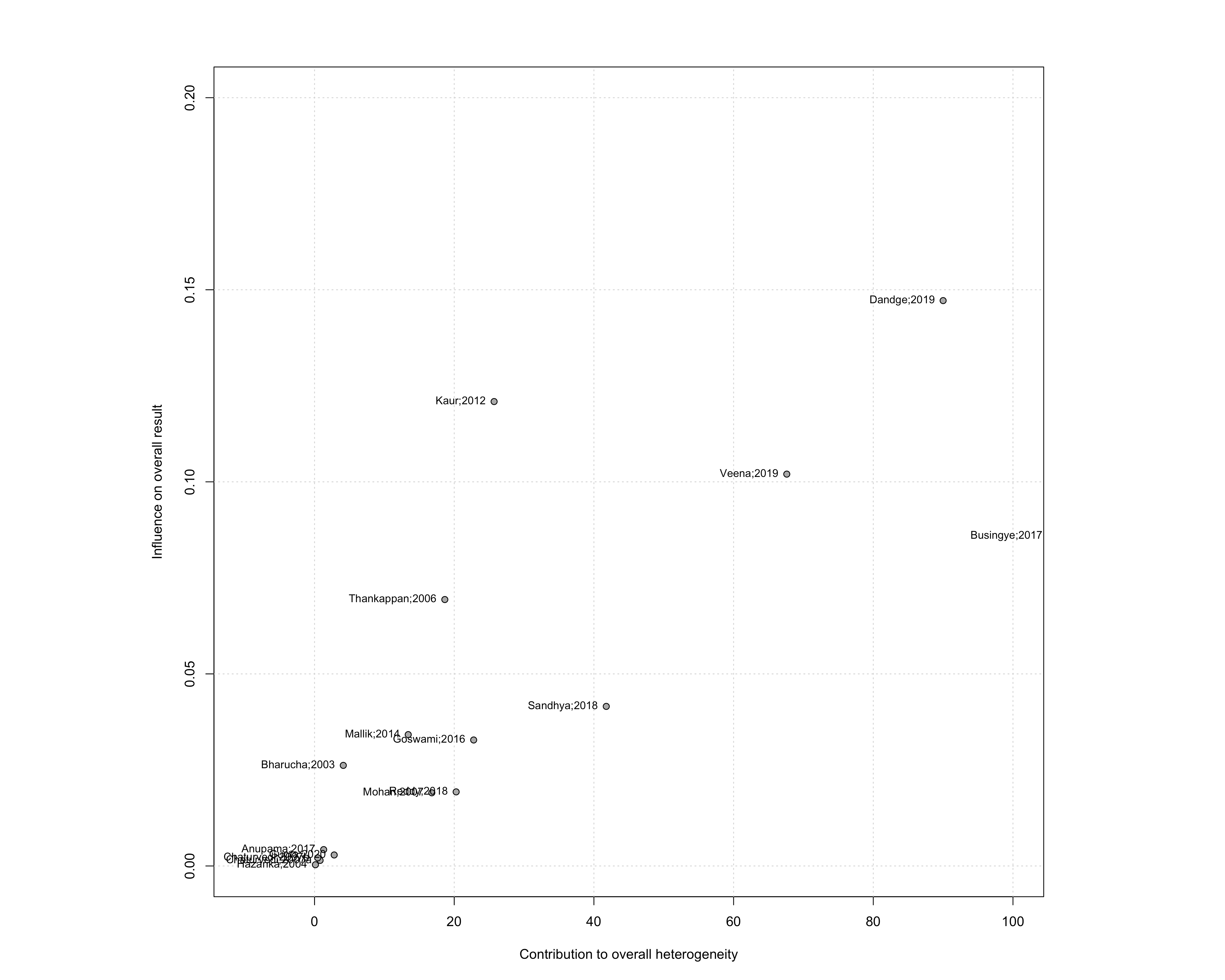

**S6: Forest plot of leave-one-out analysis**

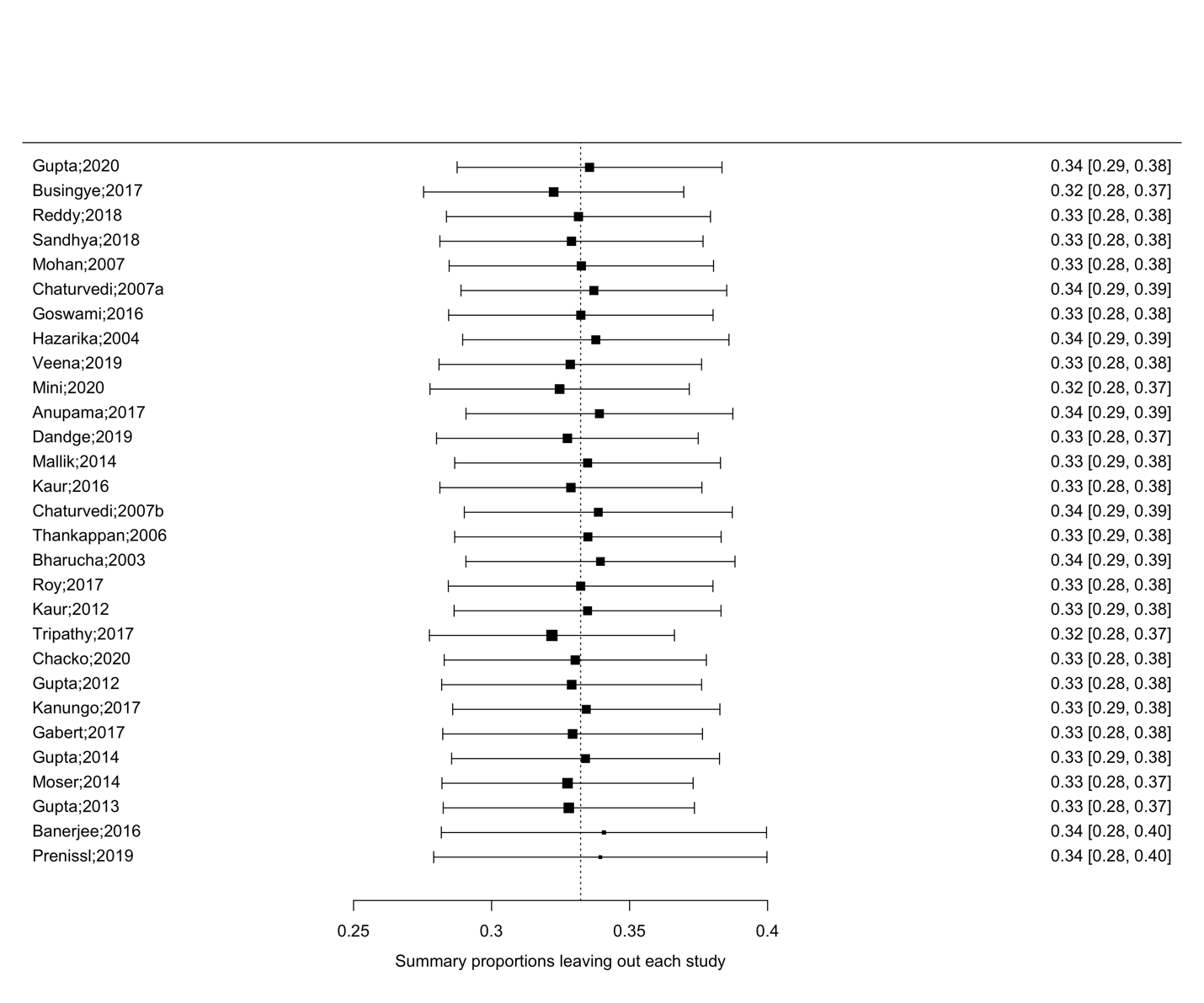

**S7: test of residual**

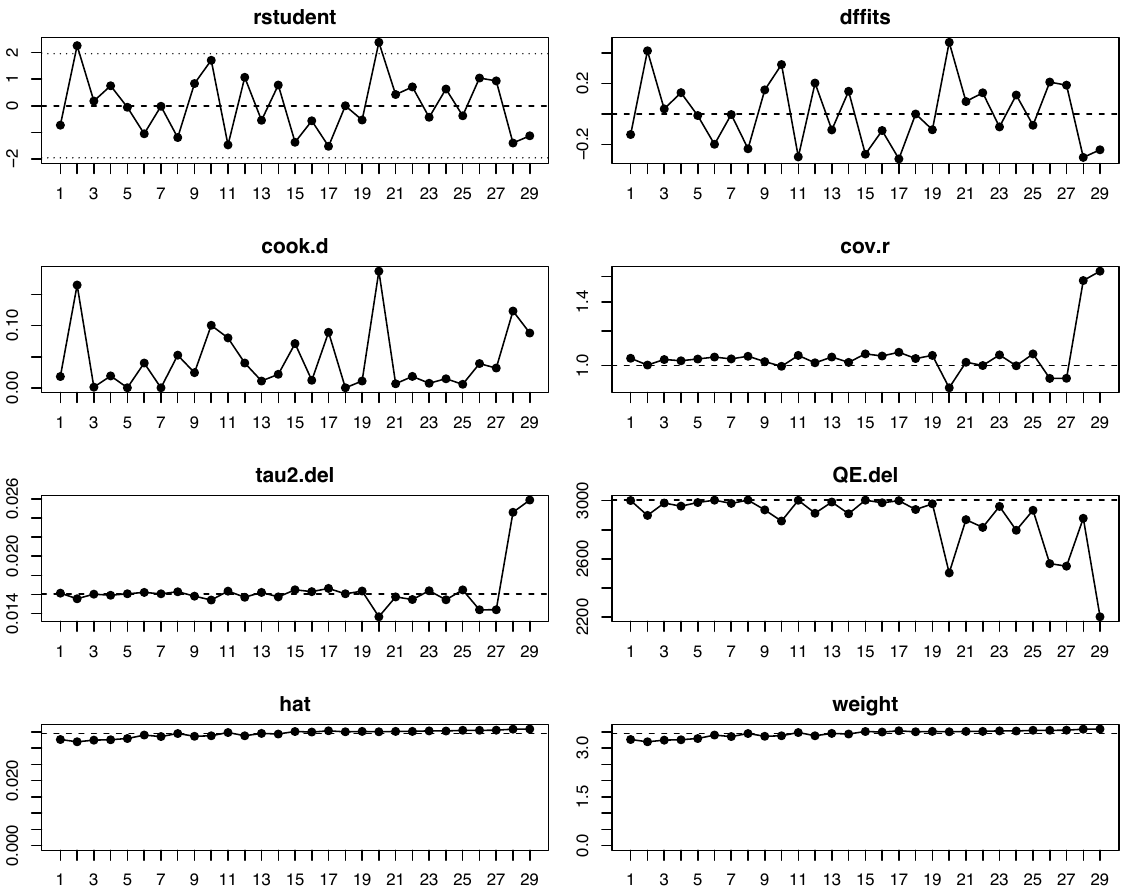

**S8: Hypertension control rates in India: sub- group analysis of studies, 2001-2020**

| **Group** | **Control rate (%),**  **[95% CI (%)]** | **SD**  **(τ, %)** | **Cochran’s Q^#^** | **p- value** |
| --- | --- | --- | --- | --- |
| **Study period** |  |  |  |  |
| 2001-2010 *n=12* | 29.6 [22.2-37.1] | 12.5 | 1.5 _df=1_ | 0.22 |
| 2011-2020 *n=17* | 35.8 [27.8-43.7] | 11.6 |  |  |
| **Regions** |  |  |  |  |
| North *n=4* | 25.3 [10.9-39.7] | 8.7 | 13.9 _df=3_ | 0.003 |
| East *n=4* | 20.7 [8.7-32.8] | 7.4 |  |  |
| South *n=13* | 39.3 [31.5-47.0] | 12.4 |  |  |
| West *n=3* | 32.9 [0-94.9] | 24.9 |  |  |
| **Sex** |  |  |  |  |
| Males *n=18* | 28.2 [21.0-35.4] | 11.4 | 1.5 _df=1_ | 0.22 |
| Females *n=20* | 34.2 [26.6-41.9] | 12.6 |  |  |

*SD- standard deviation; CI: Confidence interval; df: degree of freedom*

*n- number of studies; ^#^ Between study Q statistic*

**S9: Funnel plot**

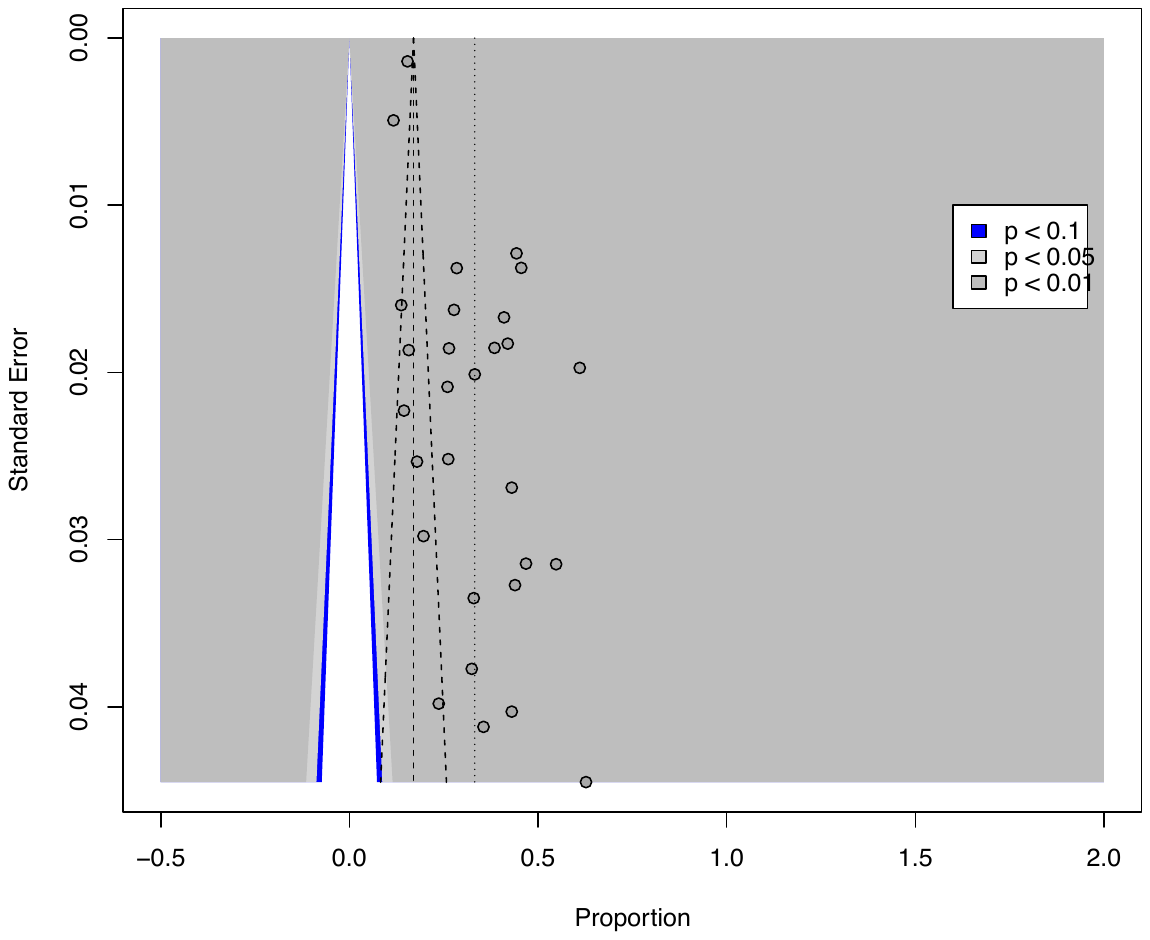
